# Routine health information system data quality in a hill and mountain district of Nepal: a cross-sectional desk review of four years of DHIS2 data from Sankhuwasabha, 2021-2025

**DOI:** 10.64898/2026.11.10.26367696

**Authors:** Dipesh Shrestha, Pramana Adhikari, Mahesh Bhatta

**Affiliations:** Health Office, Sankhuwasabha District, Khandbari, Koshi Province, Nepal; Health Section, Silichong Rural Municipality, Sankhuwasabha District, Koshi Province, Nepal

**Author notes:** Corresponding author: (DS).

## Abstract

**Introduction:** Nepal’s routine health information system runs on DHIS2, but no published data-quality assessment covers its mountainous districts. The method raises a second issue: the WHO Data Quality Review (DQR) tests internal consistency partly through related-indicator pairs, assuming each indicator is recorded independently. DHIS2 validation rules act on paired values at entry, so a rule-governed pair may agree for reasons unrelated to recording quality.

**Methods:** In this study, a WHO DQR Module 1 desk review was conducted on aggregated DHIS2 data from all 132 facilities in Sankhuwasabha district, analysed at district and local level (palika) resolution: seven datasets for completeness and timeliness, and 17 tracer indicators for internal consistency, for fiscal year 2081/82 (2024/25) against the three preceding years (2021/22 to 2023/24). Pairs were classified in advance as rule-governed or free, and three denominator-free ratios compared with Nepal Demographic and Health Survey (NDHS) 2022 estimates for Koshi Province.

**Results:** Completeness was at least 90.6% in every dataset and unit. Timeliness fell below 90% in 22 of 70 unit-dataset combinations, concentrated in four remote local levels. Of four rule-governed pairs only iron and deworming changed materially, 1.42 to 1.00; two never agreed, indicating rules that query rather than block entry. The free pairs diverged: Penta-1 to Penta-3 dropout fell from +3.6% to −5.4%, negative in 9 of 10 local levels, and BCG was below Penta-1 in 8 of 10. District totals were inverted at both steps (BCG 1,537, Penta-1 1,666, Penta-3 1,756), which redistribution cannot produce.

**Conclusion:** Reporting is nearly complete, but the data carry a persistent immunization anomaly that completeness and timeliness miss, pointing to a systematic recording process. Because a rule-governed pair may agree by configuration, DHIS2 assessments should classify pairs against the active validation-rule set before interpretation.

## Introduction

Routine health information systems generate the data that health managers use for planning, budgeting, and supervision, and their usefulness depends on how well facilities record and report the care they provide [1,2]. Nepal moved its national health management information system (HMIS) onto DHIS2 in 2016. The platform now carries monthly reports from every public health facility in the country, covering safe motherhood, immunization, family planning, nutrition, communicable disease, and outpatient care. DHIS2 serves the same role in more than 70 low- and middle-income countries, and its adoption has been accompanied by standardised methods for checking the data it holds [3,4].

The most widely used of these is the WHO Data Quality Review (DQR) [5]. Its first module measures whether expected reports arrive (completeness), whether they arrive on time (timeliness), and whether reported values are internally consistent: stable over time, free of implausible outliers, and consistent between indicators that have a predictable relationship to one another, such as the first and third doses of the same vaccine [5]. Consistency in some form appears across general frameworks of health data quality [6], and health workers in Nepal already cross-check DHIS2 figures informally at periodic review meetings [7]. Assessments built on these metrics have been published for Nepal at province and multi-country level. A desk review across the 12 districts of Lumbini Province found reporting completeness of 98 to 100% and timeliness of 94 to 96%, with internal consistency the weakest dimension [4]. A five-country analysis that reported Nepal separately found high completeness and generally consistent vaccine reporting across the onset of the COVID-19 pandemic [3]. Comparable work from Ethiopia, Tanzania, Chad, and Nigeria reports completeness that is usually, though not always, the strongest dimension, and internal consistency that is reliably the weakest, with determinants clustering around workload, training, supervision, and system design [8–11].

Two gaps motivate this study. The first is geographic: the Nepali evidence comes from the plains and mid-hill provinces, and no published data-quality assessment covers a mountainous district, where scattered settlements and difficult terrain may produce a different data-quality profile. The second is methodological and appears not to have been described previously. The related-indicator method assumes that the two indicators in a pair are recorded independently. DHIS2 configurations increasingly include validation rules that reject or query data entry when paired values disagree. A rule that rejects makes disagreement impossible in the aggregated data; a rule that queries makes it less likely. Under either mode the pair’s agreement cannot be read as independent evidence about recording behaviour, because agreement is what the rule exists to encourage. Because national validation rules and DQR pair recommendations follow the same programmatic logic, the pairs most likely to be recommended are also those most likely to be rule-governed. Where this occurs, a reported consistency result may reflect the platform’s configuration rather than the quality of recording.

This study had two aims: to assess the completeness, timeliness, and internal consistency of routine HMIS data in Sankhuwasabha district against WHO DQR benchmarks; and to examine how DHIS2 validation rules affect the behaviour of related-indicator consistency checks, using four years of data to compare pairs that a rule governs against pairs that none does.

## Materials and methods

### Study design and setting

This was a facility-based, cross-sectional desk review of secondary, aggregated DHIS2 data, with a four-year trend component. Sankhuwasabha is a hill and mountain district of Koshi Province in eastern Nepal, extending from the district headquarters at Khandbari to the Makalu Himalayan range. It had a population of 158,041 at the 2021 census and is divided into ten local levels (five municipalities and five rural municipalities), referred to here by their Nepali term, palikas. All 132 health facilities that report to DHIS2 in the district were included; there was no sampling.

Nepal uses the Bikram Sambat calendar, in which the fiscal year (FY) runs from mid-July to mid-July. The reference year was FY 2081/82, corresponding to mid-July 2024 to mid-July 2025. The three comparison years were FY 2078/79, FY 2079/80, and FY 2080/81, corresponding respectively to 2021/22, 2022/23, and 2023/24, and together spanning mid-July 2021 to mid-July 2024; Gregorian equivalents are given alongside Nepali fiscal years throughout.

### Data source and extraction

Aggregated monthly data were exported from DHIS2 by the Health Office, Sankhuwasabha, which is the district’s data custodian. Two export types were used. Reporting-rate exports provided, for each of seven datasets and each organisational unit, the expected reports, actual reports, and reports received on time. Service-volume exports provided monthly values for every data element in the seven datasets at district and palika level for all 48 months. The datasets were: Maternal and Newborn Health; Immunization; Integrated Management of Neonatal and Childhood Illnesses (IMNCI); Nutrition; Family Planning; Tuberculosis; and Outpatient Morbidity (OPD). Reporting-rate exports were made on 24 June 2026 and service-volume exports on 1 July 2026, and the analytic dataset was fixed on 1 July 2026.

### Tracer indicators

Following the DQR principle of a parsimonious tracer set, 17 indicators spanning six datasets were pre-specified for the internal-consistency analyses. S1 Table lists the exact DHIS2 data-element names and the components of every derived indicator.

Several of these indicators are not held in DHIS2 as a single data element and had to be derived by adding up the elements that make them up. Institutional delivery is the clearest example. The Sankhuwasabha DHIS2 instance contains no single “institutional delivery” element. Instead, every facility delivery is recorded under one of seven mutually exclusive elements that combine the mode of delivery with the presentation of the fetus: spontaneous delivery (cephalic, breech, or shoulder), vacuum or forceps delivery (cephalic), and caesarean section (cephalic, breech, or shoulder). Because each delivery is counted once and only once under one of these seven elements, the total number of institutional deliveries is obtained by summing all seven. Indicators reported by age band were derived in the same way: first ANC visit, for example, is the sum of the “< 20 years” and “≥ 20 years” elements.

The Outpatient Morbidity (OPD) dataset was handled asymmetrically, and this was pre-specified. Its reporting completeness and timeliness were assessed, because DHIS2 provides dataset-level reporting metadata for it, but its service volumes were not used as consistency tracers, because the dataset contains about 290 heterogeneous morbidity elements with no single well-defined tracer count.

### Data-quality metrics and benchmarks

Four groups of metrics were computed, with benchmarks taken from the DQR toolkit and the Nepali precedent study [4,5]. Report completeness (actual as a percentage of expected reports) had a target of 100%, with 90% or more considered acceptable. Timeliness (reports received by the deadline as a percentage of expected reports) had a benchmark of 90%. Consistency over time compared each palika’s ratio of the reference-year value to its three-year mean against the same ratio for the district, flagging deviations beyond ±33%. Outliers were monthly reference-year values beyond two standard deviations (moderate) or three (extreme) from the indicator’s mean, with palikas flagged when moderate outliers exceeded 5% of the 96 indicator months assessed. This threshold is weak by construction and is reported for completeness rather than as evidence: for normally distributed data about 4.6% of observations fall beyond two standard deviations, so a 5% flag sits at essentially the null expectation of the method and will separate palikas only marginally. Related-indicator pairs were expected to agree within ±10%, with directional checks for dose sequences (Penta-1 at least as large as Penta-3).

### Classification of related-indicator pairs

Eight pairs were pre-specified. Before analysis, each was matched against the district’s DHIS2 validation-rule document, which lists the indicator relationships for which a rule is defined. Four pairs matched a defined rule: iron tablets and deworming tablets for pregnant women; PCV-1 and Penta-1; diarrhoea cases and ORS-zinc treatment; and pneumonia cases and amoxicillin treatment. These were labelled rule-governed and were not used to draw conclusions about recording quality. The remaining four have no corresponding rule: first antenatal care (ANC) visits against deworming; institutional deliveries against deliveries attended by a skilled birth attendant (SBA); Penta-1 against Penta-3 (dropout); and BCG against Penta-1. Consistency conclusions rest on these free pairs. S2 Table lists every pair with its expected relationship and its rule classification.

Two limits of that document should be stated plainly, because they bound what can be concluded. It records which relationships have a rule, but neither the **enforcement mode**, that is, whether entry is blocked outright or merely queried with a warning the user may override, nor the **date each rule was enabled**. Enforcement mode and timing were therefore not known independently of the data. All eight pairs were computed for each of the four years, and the resulting series is reported below as an observation about how the pairs behaved, not as a measurement of when any rule took effect.

### External plausibility check

A full external-consistency analysis was not possible: NDHS 2022 is representative at province level, not district level, and converting facility counts to coverage would import population-projection error. Instead, three ratios that require no population denominator were computed identically from both sources: Penta-1 to Penta-3 dropout, the BCG to Penta-1 ratio, and the SBA-attended share of facility deliveries. Survey values were taken from NDHS 2022 estimates for Koshi Province [12], and facility values from FY 2078/79 (2021/22), the fiscal year overlapping the survey’s recall window. The third of these is not a like-for-like comparison and is reported with that caveat: the DHS measure is cadre-defined, counting births attended by a skilled provider wherever they occurred, including at home, whereas the facility measure is the SBA-attended share of deliveries that took place in a facility. The two denominators differ, so the comparison tests order of magnitude rather than agreement.

### Analysis

Analyses were performed in Python 3.12.0. Two third-party packages were used: xlrd 2.0.2 to read the DHIS2 .xls exports [13], and openpyxl 3.1.5 to write the analysis workbook [14]. All remaining operations used the Python standard library, principally the statistics, collections, glob, os, sys, and argparse modules. The analysis is deterministic: every metric is a descriptive computation applied to the extracted values, and the pipeline contains no simulation, resampling, or other stochastic step, so no random-number seed applies and results do not depend on the interpreter’s random number generator. Exact package versions are pinned in the lockfile distributed with the code. The code is openly available at https://github.com/Dipesh1123/dhis2-dqr-pipeline and archived on Zenodo (doi: 10.5281/zenodo.21211933), and the aggregated DHIS2 exports and the analysis workbook are deposited on Zenodo (doi: 10.5281/zenodo.23149871). The scripts read the DHIS2 exports and write all metrics to a single analysis workbook, which reproduces every value reported in this paper from the raw exports.

The aggregated exports were analysed as extracted, with no imputation; blank monthly cells were treated as zero. Reference-year monthly series for all tracer indicators were checked for gaps before analysis, and none was found. Two safeguards against interpretation bias were built in. First, all benchmarks, tracer indicators, and the pair classification were specified before any metric was computed. Second, indicators with small absolute counts, namely tuberculosis with 76 notified cases district-wide in the reference year, were flagged in advance for cautious interpretation, because small denominators inflate ratio deviations. Results are presented at district and palika resolution, with a composite report card per palika.

The study is reported in accordance with the Strengthening the Reporting of Observational Studies in Epidemiology (STROBE) statement [15] and its RECORD extension for studies using routinely collected health data [16]. The completed checklist is provided as S1 Checklist.

### Sex and gender

The tracer indicators were extracted as aggregated facility totals and were not disaggregated by sex in this analysis. The maternal indicators, namely antenatal care, institutional delivery, and skilled birth attendance, relate to women by definition, and the immunization, child health, nutrition, and tuberculosis tracers were analysed as combined counts. Because the study assesses the quality of the reporting system rather than any health outcome, no metric is interpreted by sex or gender. This is reported in line with the Sex and Gender Equity in Research (SAGER) guidelines, which ask that the absence of sex and gender reporting be explained.

### Ethics statement

The study used only aggregated, de-identified administrative data with no human participants, and therefore falls outside the scope of human-subject research as defined in the Nepal Health Research Council’s national ethical guidelines; the same classification was applied in a comparable published desk review [4]. For this reason ethics committee review was not sought, and no consent procedure applied. Administrative authorization to use the data for research was obtained from the Health Office, Sankhuwasabha, the district data custodian. The data were accessed for research purposes on 24 June 2026 (reporting rates) and 1 July 2026 (service volumes). The exports contain only aggregated monthly counts and reporting rates, with no individual-level records, and the authors had no access to information that could identify individual patients during or after data collection.

### Use of artificial intelligence tools

Claude (Anthropic), a large language model, was used under the authors’ direction to assist with writing the Python analysis scripts and with drafting, editing, and formatting the manuscript. It was not used to generate, alter, or impute data. The authors evaluated its outputs by verifying all data, computations, references, and interpretations against the DHIS2 exports, the analysis workbook, and the original sources, and take full responsibility for the content.

## Results

### Completeness and timeliness

Report completeness was uniformly high. Across seven datasets and eleven reporting units (district plus ten palikas), completeness never fell below 90.6%, and most unit-dataset combinations reached 100%. At district level, completeness ranged from 99.0% (Nutrition) to 100% (Immunization and IMNCI) (Table 1). No reporting unit was excluded and no filtering on data availability was applied.

**Table 1.** District-level report completeness and timeliness by dataset, FY 2081/82 (2024/25).

| **Dataset** | **Completeness (%)** | **Timeliness (%)** |
|---|---|---|
| Immunization | 100.0 | 91.2 |
| IMNCI | 100.0 | 96.0 |
| Nutrition | 99.0 | **88.9** |
| Maternal and Newborn Health | 99.4 | **89.7** |
| Family Planning | 99.8 | 93.0 |
| Tuberculosis | 99.4 | 91.5 |
| Outpatient Morbidity | 99.6 | 96.9 |

*Bold values fall below the 90% timeliness benchmark. OPD was assessed for reporting performance only (see Methods).*

Timeliness was the weaker dimension and showed a clear geographic pattern. Two datasets missed the 90% benchmark at district level, Nutrition (88.9%) and Maternal and Newborn Health (89.7%), and 22 of 70 palika-dataset combinations fell below 90%. Late reporting concentrated in four of the larger and more remote units: Bhotkhola, Chainapur, Dharmadevi, and Khandabari. The lowest values were in Nutrition at Chainapur (69.9%) and Bhotkhola (72.7%). Makalu, Silichong, Madi, and Panchakhapan were consistently timely.

### Related-indicator pairs: rule-governed pairs

The eight related-indicator pairs divided along the pre-specified classification. In the reference year, all four rule-governed pairs returned ratios of about 1.00 in nearly every palika, with only two isolated deviations, both of 11% or less.

The four-year series (Table 2, Fig 1A) shows that the four rule-governed pairs did not behave alike, and the differences between them matter for interpretation. Only one pair, iron and deworming, changed substantially: it stood at 1.42 in FY 2078/79 (2021/22), a 42% difference of the kind the related-indicator method is designed to detect, and moved to 1.03, then to 1.00 in each of the two most recent years. Penta-1 and PCV-1 sat at 1.00 in all four years, including the baseline year, so it shows no change and carries no information about whether or when anything was enforced. Diarrhoea and ORS-zinc narrowed but never reached agreement (1.11, 1.07, 1.04, 1.03), and pneumonia and amoxicillin moved without direction, falling to 0.98 before returning to 1.00.

**Fig 1.**
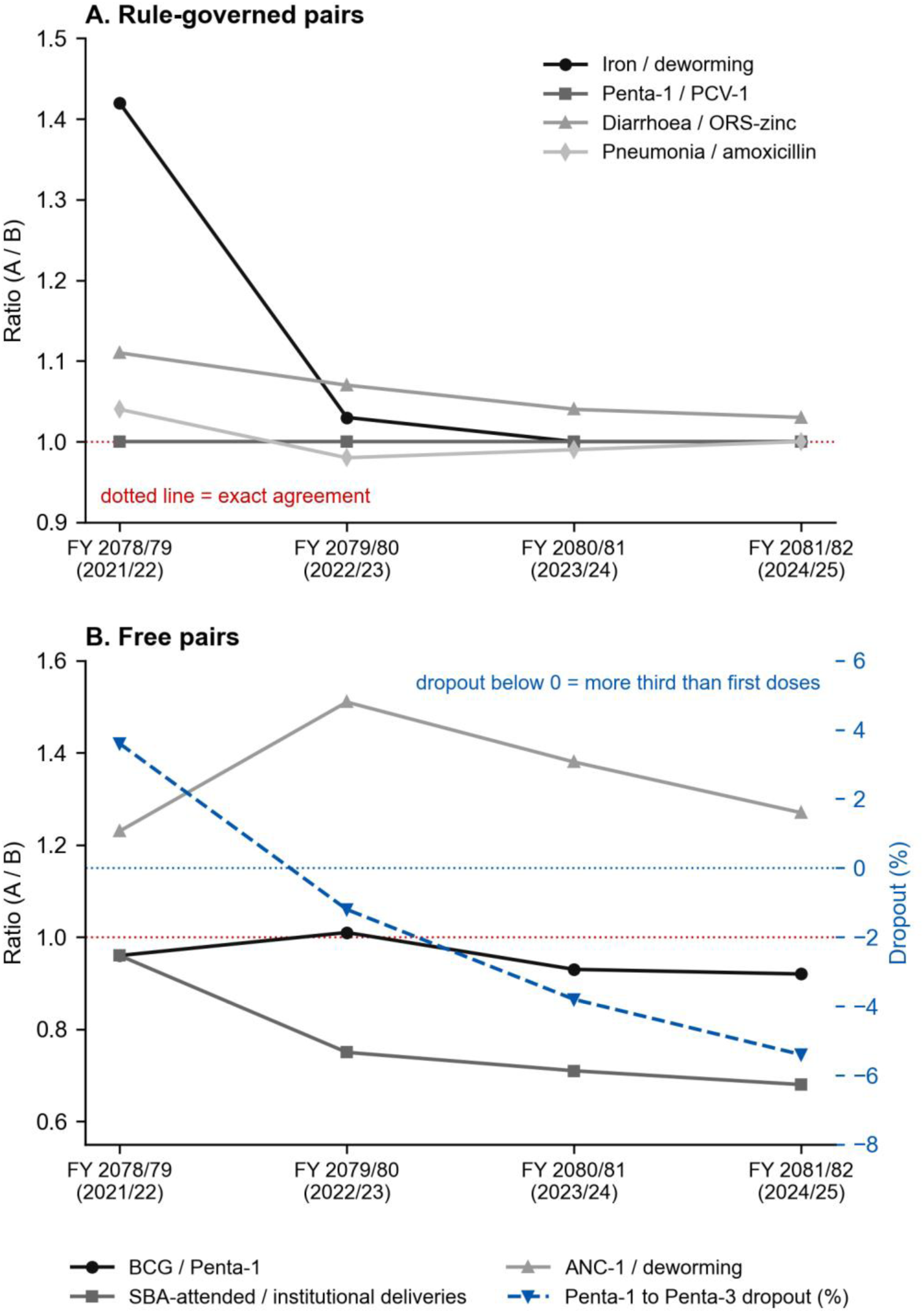
District-level related-indicator pair metrics across four fiscal years, FY 2078/79 (2021/22) to FY 2081/82 (2024/25). (A) The four rule-governed pairs: only iron and deworming moves, from 1.42 to agreement, while diarrhoea and ORS-zinc never reach the dotted line of exact agreement and pneumonia and amoxicillin cross below it. (B) The four free pairs, with the Penta-1 to Penta-3 dropout on the right-hand axis crossing from positive to negative and continuing to fall. Values are district totals; the counts behind them are in S3 Table.

**Table 2.** District-level pair metrics across four fiscal years.

| Pair | Relationship | FY 2078/79 (2021/22) | FY 2079/80 (2022/23) | FY 2080/81 (2023/24) | FY 2081/82 (2024/25) |
|---|---|---|---|---|---|
| Rule-governed | Iron 1st time / deworming | **1.42** | 1.03 | 1.00 | 1.00 |
| Rule-governed | Penta-1 / PCV-1 | 1.00 | 1.00 | 1.00 | 1.00 |
| Rule-governed | Diarrhoea / ORS-zinc | 1.11 | 1.07 | 1.04 | 1.03 |
| Rule-governed | Pneumonia / amoxicillin | 1.04 | 0.98 | 0.99 | 1.00 |
| Free | ANC-1 / deworming | 1.23 | 1.51 | 1.38 | 1.27 |
| Free | SBA-attended / institutional deliveries | 0.96 | 0.75 | 0.71 | 0.68 |
| Free | Penta-1 to Penta-3 dropout (%) | **+3.6** | **−1.2** | **−3.8** | **−5.4** |
| Free | BCG / Penta-1 | 0.96 | 1.01 | 0.93 | 0.92 |

Negative dropout, meaning more third doses than first doses, cannot occur in a true cohort. Ratios are shown to two decimal places; pairs displayed as 1.00 are not necessarily equal, and the underlying counts are given in S3 Table.

Two of these observations bear directly on how strong a claim the data support. First, a rule that blocked unequal entry outright would force a ratio of exactly 1.00, yet diarrhoea and ORS-zinc remained 3% apart in the reference year and pneumonia and amoxicillin fell below 1.00 in FY 2079/80. Neither is compatible with a hard constraint. Second, the pairs that display as 1.00 are not in fact equal: in the reference year iron was 1,500 against deworming 1,499, and PCV-1 was 1,667 against Penta-1 1,666. Each differs by a single record and rounds to 1.00 at two decimal places. The data therefore show **convergence in one pair**, not demonstrable enforcement in four, and are most consistent with rules that query rather than block. What follows for interpretation is narrower than a tautology but still consequential: where a rule governs a pair, its agreement cannot be read as independent evidence about recording, because agreement is what the rule exists to encourage.

### Related-indicator pairs: free pairs

The four pairs not governed by a validation rule carried the informative findings, and their divergence over the four years is shown in Fig 1B.

Penta dropout was negative and worsened in each successive year. District dropout moved from +3.6% in FY 2078/79 (2021/22) to −1.2%, −3.8%, and −5.4% in the following three years. In the reference year, Penta-3 doses (1,756) exceeded Penta-1 doses (1,666) in 9 of 10 palikas; only Panchakhapan recorded a normal positive dropout (+9.2%). The largest negative values were in Chichila (−35.2%) and Makalu (−12.4%), and seven palikas recorded negative dropout in at least two of the four years.

BCG counts were persistently lower than Penta-1. BCG, given at birth, would be expected to be at least as frequent as Penta-1, but it was lower in 8 of 10 palikas in the reference year (ratios 0.76 to 0.92). Silichong and Chichila were below 0.90 in all four years. The consistent exception was Khandabari, the headquarters municipality that hosts the district hospital’s birthing centre, where BCG exceeded Penta-1 in every year (1.15 to 1.24). The two patterns are complementary: a surplus of birth doses in the municipality with the birthing centre, and a deficit in the others.

The SBA-attended share of institutional deliveries fell steadily at district level, from 0.96 to 0.68 over the four years, and ranged from 0.14 to 1.00 across palikas in the reference year. In Bhotkhola, 10 of 69 facility deliveries were recorded as SBA-attended. Variation of this magnitude in a subset indicator is difficult to explain by service patterns alone.

The remaining free pair behaved unremarkably: ANC-1 exceeded deworming by 23 to 51% in every year. Because the national protocol schedules deworming at a second-trimester contact rather than at the first ANC visit, this gap is plausibly explained by the timing of care rather than by a recording error.

### External plausibility check

In FY 2078/79 (2021/22), the year overlapping the NDHS 2022 recall window, facility ratios were consistent with the independent survey on all three denominator-free checks (Table 3). The anomalous values appeared only in later years, for which no survey benchmark exists.

**Table 3.** Denominator-free ratios: NDHS 2022 (Koshi Province) against facility data (Sankhuwasabha).

| **Metric** | **NDHS 2022 Koshi** | **Facility FY 2078/79 (2021/22)** | **Facility FY 2081/82 (2024/25)** |
|---|---|---|---|
| Penta-1 to Penta-3 dropout | +1.4% | +3.6% | **−5.4%** |
| BCG / Penta-1 | 1.01 | 0.96 | 0.92 |
| SBA-attended / facility deliveries | 1.00 | 0.96 | **0.68** |

Survey estimates: BCG 94.3%, Penta-1 93.2%, Penta-3 91.9% (children 12-23 months, n=168); facility delivery 81.5%, skilled attendance 81.8% (births in the two preceding years, n=368) [12].

### Consistency over time, outliers, and the composite report card

Ten palika-indicator combinations breached the ±33% consistency-over-time band, but six of the ten were tuberculosis notifications, an indicator with only 76 district-wide cases in the reference year, for which a small number of cases moves the ratio outside any threshold. Excluding tuberculosis, over-time flags were isolated. Outlier loading was low: only Sabhapokhari (6.2%) and Dharmadevi (5.2%) exceeded the 5% threshold for moderate outliers, and the whole reference year contained four extreme outliers. Service volumes contracted across the four years (ANC-1 by 29%, institutional deliveries by 26%, new family-planning acceptors by 42%), consistent with the district’s declining and out-migrating population; tuberculosis notification was the only tracer to rise (+19%).

Combining the dimensions, five palikas were classified as priority for data-quality support (Bhotkhola, Makalu, Chichila, Sabhapokhari, Dharmadevi) and five for routine review. Flags were driven by timeliness and immunization consistency rather than by completeness.

## Discussion

### Principal findings

Four years of routine data from one hill and mountain district produced three main findings. First, the district’s reporting is almost complete but not uniformly timely, and lateness clusters in a small number of remote palikas. Second, its immunization dose counts carry a persistent and worsening internal-consistency anomaly that is ordered across the dose sequence and survives aggregation to district level, so it reflects a systematic recording process rather than movement of infants between palikas. Third, and with the widest relevance beyond this district, half of the pre-specified consistency checks were governed by validation rules, and the four-year behaviour of those pairs shows that a rule shapes what such a check can report.

### Validation rules and the consistency toolkit

The related-indicator method assumes independent recording. Where a validation rule acts on the pair, that assumption is at least weakened and may fail entirely. The four-year series shows the weakened case rather than the extreme one. Iron and deworming differed by 42% in the baseline year and agreed to within a single record thereafter, so its agreement in the reference year says little about how facilities record either service. Two other governed pairs never agreed at all, which indicates that these rules query entry rather than block it, and a third sat at agreement in every year including the baseline and so carries no information either way. The honest reading is that a governed pair cannot be treated as independent evidence about recording, not that the check has been rendered incapable of failing. Had this assessment covered only the reference year, it would still have reported four pairs in close agreement and credited the district with a consistency it had not demonstrated, which is the practical risk.

This interaction is not specific to Sankhuwasabha. Validation rules and DQR pair recommendations derive from the same programmatic logic, so the indicator pairs most likely to be recommended for consistency checking are also those most likely to be rule-governed. Neither the Nepali province-level assessment nor the multi-country DHIS2 analysis distinguished rule-governed from free pairs [3,4], and the DQR toolkit does not require the distinction [5]. A straightforward amendment would address this: classify each candidate pair against the active validation-rule set before analysis; report rule-governed pairs only as evidence that rules are being applied at data entry; and base consistency conclusions on free pairs, using pre-enforcement years to recover information where historical data are available. Publishing national validation-rule configurations alongside data exports would allow any assessment to make this classification without insider access to the platform. Publishing the enforcement mode alongside the rule, that is, whether entry is blocked or merely queried, would settle how much weight a governed pair can bear; the district document available here records the relationships but not the mode, which is why this study can report convergence but not enforcement.

### The immunization anomaly

Once the rule-governed pairs are set aside, the informative checks converge on immunization, and the pattern is an ordered one. In the reference year the district recorded 1,537 BCG doses, 1,666 Penta-1 doses and 1,756 Penta-3 doses. The sequence is inverted at both steps: fewer doses at each earlier contact than at the one that follows it, when a true birth cohort can only lose members as it moves through the schedule.

An earlier reading of these data attributed the pattern to catchment misattribution, on the basis that Khandabari, the headquarters municipality hosting the district hospital’s birthing centre, is the one palika where BCG exceeds Penta-1. That mechanism cannot account for what is observed, and the arithmetic is worth setting out because it constrains the explanation. Movement of infants between palikas redistributes counts; it cannot change the district total. Khandabari’s surplus of BCG over Penta-1 is 64 doses, while the other nine palikas together show a deficit of 193, leaving a net district deficit of 129. The dropout gap behaves the same way, with 90 more third doses than first doses district-wide. Redistribution between palikas cannot produce either figure. Nor does catchment address the dropout inversion at all, since it concerns where a birth dose is recorded rather than why later doses outnumber earlier ones.

What the data indicate is therefore a single systematic process acting across the district, not two local ones. Three candidates are consistent with an ordered inversion of this kind and cannot be separated without source records: under-recording or under-reporting of the birth dose relative to later contacts; counting of later doses against sessions, vials or opportunities rather than against individual children; and register or tally practices that carry children forward without matching them back to a first dose. Comparable failures of expected numerical relationships in DHIS2 maternal and newborn indicators have been reported from Nigeria [11], and the upstream register-quality problems that could produce them are well described [17]. Catchment movement may still contribute to the geographic spread, and specifically to Khandabari being the sole palika above 1.00, but it is not the explanation for the district-level result and is no longer offered as one.

The external check is consistent with a change in recording rather than in the population: facility ratios agreed with NDHS 2022 in the one year the survey covered and diverged afterwards, and a change of this size over this period is more readily explained by recording practice than by demography. The practical consequence does not depend on identifying the mechanism. Dropout rate, a core immunization management indicator, is currently uninterpretable in this district at either palika or district level, and any coverage estimate built on these dose counts inherits the same distortion [18]. None of this was visible in the completeness or timeliness metrics, which is the main argument this study offers for routine internal-consistency checking at district level.

### Timeliness, small counts, and shrinking denominators

The contrast between near-universal completeness and patchy timeliness localises the reporting problem: reports arrive, but late, and late reporting concentrates where terrain and workload are most difficult. This is addressable through targeted support to four named palikas rather than a district-wide intervention. Three interpretive cautions apply. Tuberculosis, with 76 cases a year, generated most of the over-time flags through small-count volatility alone, and a minimum-count filter would prevent such false alarms. The outlier dimension separated almost nothing, which is what its threshold predicts: flagging palikas whose moderate outliers exceed 5% of assessed indicator months sets the bar at the null expectation for normally distributed data, so the two palikas that crossed it, at 6.2% and 5.2%, are barely distinguishable from noise and should not carry weight in the composite report card. And because the district’s service volumes are contracting with its population, trend-based checks will flag declines that are demographic rather than data-quality in origin, which is a further reason to anchor district-level assessment in internal-consistency logic.

### Strengths and limitations

This assessment drew on reports from every facility in the district rather than a sample, applied benchmarks and a pair classification specified before any metric was computed, and derived every indicator from named DHIS2 elements in a workbook that reproduces each reported value. The four-year series shows behaviour that single-year assessments cannot observe, and the baseline year was corroborated against an independent survey using ratios that avoid population-denominator error.

The limitations are substantial and bound the conclusions closely. Although all 132 facilities report into the datasets analysed, every metric is computed at district or palika level, giving ten local units and 70 unit-dataset combinations. The analysis is facility-based in its source but not facility-level in its resolution, so heterogeneity between facilities within a palika is invisible here, and a facility-level assessment of a mountainous district remains to be done. The district’s validation-rule document records which relationships carry a rule but neither the enforcement mode nor the date each was enabled, so the four-year series is an observation about pair behaviour and not a measurement of enforcement. That two rule-governed pairs never reached agreement suggests these rules query rather than block, which weakens any claim that a check was rendered incapable of failing. Confirming this requires the DHIS2 configuration log and the rule definitions themselves. Ratios computed on small denominators move sharply for few events; Chichila’s dropout of −35.2% rests on 54 first doses and 73 third doses, a difference of 19. S3 Table gives the counts behind every palika ratio so readers can weigh them, and a minimum-count filter would be a sensible addition to routine use. The systematic processes proposed for the immunization inversion are candidates, not findings, and separating them requires source-record verification the study did not perform. The check is a plausibility comparison, not a formal external-consistency analysis: NDHS 2022 is representative at province level, its immunization estimates rest on 168 children, and no survey covers the years in which the anomalies emerged. The skilled-attendance comparison is additionally a construct mismatch and is interpreted as such (see Methods). Reference-year monthly series were checked for gaps and none was found, but that check was not repeated for the three comparison years on which the four-year series depends, so blank cells there are treated as zero without verification. Finally, the palika-level findings belong to a single district, although the caution about rule-governed pairs applies wherever DHIS2 validation rules are active, and longitudinal measurement of this kind has proved feasible in other Nepali surveillance settings [19].

## Conclusions

Routine health data in Sankhuwasabha are nearly complete and partially late, and they carry a persistent immunization recording anomaly that completeness monitoring does not detect. The district totals are inverted at both steps of the dose sequence by margins that movement between local levels cannot produce, which points to a systematic recording process that source-record verification will be needed to identify. The four-year record also shows how validation rules shape what the WHO DQR’s related-indicator checks can report: one governed pair converged from a 42% difference to agreement, while two others never agreed, indicating rules that query rather than block. As validation-rule coverage expands across DHIS2 implementations, assessments will need to distinguish agreement that reflects recording from agreement the platform encourages, and to say which of the two a reported result represents.

## Supporting information

Supporting Information (S1-S3 Tables, S1 Text, S1 Checklist)

## Funding

The authors received no specific funding for this work.

## Competing interests

All authors are employed within the Sankhuwasabha district health system whose reporting is assessed in this study, and DS and MB are employed by the Health Office, Sankhuwasabha, which is the custodian of the data analysed. The authors declare no financial interests.

## Data availability

The aggregated DHIS2 exports (reporting rates and monthly service volumes for Sankhuwasabha district, FY 2078/79 to 2081/82) and the analysis workbook that reproduces every reported value are available from the Zenodo repository under a CC BY 4.0 licence (doi: 10.5281/zenodo.23149871). The analysis code is available at https://github.com/Dipesh1123/dhis2-dqr-pipeline and archived on Zenodo (doi: 10.5281/zenodo.21211933). S3 Table gives the counts behind every palika-level ratio.

## Author contributions

**Dipesh Shrestha:** Conceptualization, Methodology, Data curation, Formal analysis, Writing (original draft), Writing (review and editing).

**Pramana Adhikari:** Writing (original draft), Writing (review and editing).

**Mahesh Bhatta:** Supervision, Methodology, Writing (review and editing).

## Supporting information

**S1 Table. Tracer indicators: exact DHIS2 data-element names, derivations, and metric assignments.**

**S2 Table. Related-indicator pairs and validation-rule classification.**

**S3 Table. Counts behind every palika ratio, reference year FY 2081/82 (2024/25).** Numerators and denominators for the immunization and delivery ratios by local level, and the district aggregation check for the immunization pattern.

**S1 Text. Reproducibility.** Computing environment, software versions, and the steps needed to reproduce every reported value from the DHIS2 exports.

**S1 Checklist. STROBE checklist with the RECORD extension for studies using routinely collected health data.**

## Notes

### Author Declarations

The study used only aggregated monthly counts and reporting rates from the DHIS2 routine health information system, with no individual-level records and no human participants. All data analysed are openly available at https://doi.org/10.5281/zenodo.23149871 under a CC BY 4.0 licence. Under the Nepal Health Research Council's national ethical guidelines such work falls outside the scope of human-subject research, so ethics committee review was not sought. Administrative authorization to use the data for research was obtained from the Health Office, Sankhuwasabha, the district data custodian.

