## Supporting Information (S1-S3 Tables, S1 Text, S1 Checklist) for "Routine health information system data quality in a hill and mountain district of Nepal: a cross-sectional desk review of four years of DHIS2 data from Sankhuwasabha, 2021-2025"

#### S1 Table. Tracer indicators: exact DHIS2 data-element names, derivations, and metric assignments

T = consistency over time; O = outlier analysis; R = related-indicator pair.

Derived indicators are computed as the sum of the listed component elements. Element names are reproduced exactly as they appear in the DHIS2 exports for Sankhuwasabha district.

| S.N. | **Indicator (exact DHIS2 element name or derivation)** | **Dataset** | **T** | **O** | **R** |
|---|---|---|---|---|---|
| 1 | Safe Motherhood Program-Antenatal Checkup-First ANC Visit (any time): sum of "< 20 Years" + "≥ 20 Years" | Maternal and Newborn Health (07) | x | x | x |
| 2 | Institutional delivery: sum of the 7 "Safe Motherhood Program-Type of Delivery-" elements, namely Spontaneous Cephalic, Spontaneous Breech, Spontaneous Shoulder, Vaccum/Forcep Cephalic, C/S Cephalic, C/S Breech, and C/S Shoulder. There is no single "institutional delivery" element in this DHIS2 instance; each facility delivery is recorded once under exactly one of these seven mutually exclusive elements, so their sum is the total. | Maternal and Newborn Health (07) | x | x | x |
| 3 | MNH-Delivery Service-Skilled Birth Attendants (SBA)Facility | Maternal and Newborn Health (07) | | | x |
| 4 | Immunization Program-Vaccine Type-Children Immunized-BCG Doses | Immunization (02-1) | x | x | x |
| 5 | Immunization Program-Vaccine Type-Dose-Children Immunized-DPT-HepB-Hib-1st (Penta-1) | Immunization (02-1) | x | x | x |
| 6 | Immunization Program-Vaccine Type-Dose-Children Immunized-DPT-HepB-Hib-3rd (Penta-3) | Immunization (02-1) | | | x |
| 7 | Immunization Program-Vaccine Type-Dose-Children Immunized-PCV-1st | Immunization (02-1) | | | x |
| 8 | Immunization Program-Children Immunized-Measles/Rubella-9-11 Months | Immunization (02-1) | x | x | |

| | | | | | |
|---|---|---|---|---|---|
| 9 | New family-planning acceptors: sum of "Family Planning Program-Temporary FP Method-{Pills, Depo, IUCD, Implant (5 Years)}-New Users {<20, ≥20} Years" (8 elements) | Family Planning (08) | x | x | |
| 10 | Nutrition-Pregnant Women Receiving-Iron Tablets at 1st time | Nutrition (03-1) | | | x |
| 11 | Nutrition-Pregnant Women Receiving-Deworming Tablets | Nutrition (03-1) | | | x |
| 12 | Growth monitoring 0-11 months: sum of "Nutrition-Registered for GM-New Visit 0-11 Months - {Normal, Moderate, Severe}" | Nutrition (03-1) | x | x | |
| 13 | Diarrhoea cases 2-59 months: sum of "CBIMCI-(2-59Months)-Classification-Diarrhoea-{No Dehydration, Some Dehydration, Severe Dehydration, Dysentery, Prolonged Diarrhoea}" | IMNCI (02-2) | x | | x |
| 14 | CBIMCI-(2-59Months)-Treatment-ORS & Zinc | IMNCI (02-2) | | | x |
| 15 | CBIMCI-(2-59Months)-Classification-ARI-Pneumonia | IMNCI (02-2) | | | x |
| 16 | CBIMCI-(2-59Months)-Treatment-Amoxicillin | IMNCI (02-2) | | | x |
| 17 | Tuberculosis notified: sum of "TB-Age group-All New" + "TB-Age group-All Relapse" | Tuberculosis (15-1) | x | x | |

Summary: 17 tracer indicators; 9 assessed for consistency over time; 8 for outliers (outlier denominator per palika = 8 indicators x 12 months = 96); 8 related-indicator pairs (S2 Table). The Outpatient Morbidity dataset (18) was assessed for reporting completeness and timeliness only; it contains approximately 290 heterogeneous morbidity elements with no single well-defined tracer count, so no OPD volume was used in the consistency analyses (pre-specified).

#### S2 Table. Related-indicator pairs and validation-rule classification

Pairs were classified against the district's active DHIS2 validation-rule configuration before analysis. Rule-governed pairs are not used to draw conclusions about recording quality; see the manuscript Results for what the four-year series does and does not establish about rule compliance; consistency conclusions rest on the free pairs.

| Pair | Indicator A | Indicator B | Expected relationship | Classification |
|---|---|---|---|---|
| P1 | ANC first visit (any time) | Deworming tablets (pregnant women) | A ≈ B (deworming scheduled at second-trimester contact) | Free |
| P2 | Iron tablets at 1st time | Deworming tablets (pregnant women) | A = B | Rule-governed |
| P3 | Institutional deliveries | SBA-attended deliveries (facility) | B ≤ A; B/A stable across palikas | Free |
| P4 | Penta-1 | Penta-3 | A ≥ B; dropout 0-10% | Free |
| P5 | Penta-1 | PCV-1 | A = B (co-administered) | Rule-governed |
| P6 | BCG | Penta-1 | A ≥ B approximately (both reflect the birth cohort) | Free |
| P7 | Diarrhoea cases (2-59 m) | ORS and zinc treated | A = B | Rule-governed |
| P8 | Pneumonia cases (2-59 m) | Amoxicillin treated | A ≈ B | Rule-governed |

#### S3 Table. Counts behind every palika ratio, reference year FY 2081/82 (2024/25)

Ratios in the manuscript are shown to two decimal places. This table gives the numerators and denominators so readers can judge how much weight each ratio carries. Two points follow directly from it. First, pairs that display as 1.00 are not necessarily equal: district iron was 1,500 against deworming 1,499, and PCV-1 was 1,667 against Penta-1 1,666, each a difference of one record. Second, several palika ratios rest on small counts, most notably Chichila's dropout of -35.2%, which is 54 first doses against 73 third doses, a difference of 19.

**Penta-1 to Penta-3 (dropout) and BCG to Penta-1, by local level.**

| Local level | Penta-1 | Penta-3 | Dropout (%) | BCG | BCG / Penta-1 |
|---|---|---|---|---|---|
| District (Sankhuwasabha) | 1,666 | 1,756 | -5.4 | 1,537 | 0.92 |
| Bhotkhola RM | 87 | 90 | -3.4 | 75 | 0.86 |
| Makalu RM | 153 | 172 | -12.4 | 123 | 0.80 |
| Silichong RM | 88 | 93 | -5.7 | 70 | 0.80 |
| Chichila RM | 54 | 73 | -35.2 | 44 | 0.81 |
| Sabhapokhari RM | 106 | 113 | -6.6 | 97 | 0.92 |
| Khandabari M | 425 | 444 | -4.5 | 489 | 1.15 |
| Panchakhapan M | 174 | 158 | +9.2 | 155 | 0.89 |
| Chainapur M | 308 | 319 | -3.6 | 267 | 0.87 |
| Madi M | 117 | 126 | -7.7 | 89 | 0.76 |
| Dharmadevi M | 154 | 168 | -9.1 | 128 | 0.83 |

RM = rural municipality; M = municipality. Dropout is (Penta-1 minus Penta-3) divided by Penta-1.

**Institutional deliveries and SBA-attended deliveries, by local level.**

| Local level | Institutional deliveries | SBA-attended | Ratio |
|---|---|---|---|
| District (Sankhuwasabha) | 1,476 | 1,010 | 0.68 |
| Bhotkhola RM | 69 | 10 | 0.14 |
| Makalu RM | 76 | 66 | 0.87 |
| Silichong RM | 59 | 59 | 1.00 |
| Chichila RM | 18 | 12 | 0.67 |
| Sabhapokhari RM | 42 | 25 | 0.60 |
| Khandabari M | 896 | 552 | 0.62 |
| Panchakhapan M | 62 | 56 | 0.90 |
| Chainapur M | 153 | 131 | 0.86 |
| Madi M | 52 | 52 | 1.00 |
| Dharmadevi M | 49 | 47 | 0.96 |

**District aggregation check for the immunization pattern.** Khandabari records 64 more BCG doses than Penta-1 doses (489 against 425). The other nine local levels together record 193 fewer (1,048 against 1,241). The district is therefore 129 doses short of Penta-1 on BCG, and records 90 more Penta-3 than Penta-1 doses. Movement of infants between local levels redistributes counts and cannot change a district total, so neither figure can be produced by catchment misattribution.

#### S1 Text. Reproducibility

All metrics were computed by open Python scripts from the raw DHIS2 exports. The analysis was run under Python 3.12.0 with two third-party packages: xlrd 2.0.2 (reading the DHIS2 `.xls` exports) and openpyxl 3.1.5 (writing the analysis workbook). Everything else uses the Python standard library (`statistics`, `collections`, `glob`, `os`, `sys`, `argparse`). Exact versions are pinned in the `uv.lock` file distributed with the code. The pipeline is deterministic: it contains no simulation, resampling, or other stochastic step, so no random-number seed applies and the output does not depend on the interpreter's random number generator. (The only seeded random draw anywhere in the repository is in `make_synthetic_data.py`, which generates the synthetic demonstration dataset and takes no part in the analysis reported here.) The analysis workbook contains one sheet per data-quality dimension (completeness and timeliness; consistency over time; outliers; related-indicator pairs; composite report card; four-year trends) and reproduces every value reported in the manuscript. The scripts, together with a synthetic demonstration dataset that runs the full pipeline end to end, are openly available at https://github.com/Dipesh1123/dhis2-dqr-pipeline and archived on Zenodo (https://doi.org/10.5281/zenodo.21211933). The aggregated DHIS2 exports and the analysis workbook are deposited on Zenodo (https://doi.org/10.5281/zenodo.23149871), with the authorization of the data custodian, the Health Office, Sankhuwasabha.

### S1 Checklist. STROBE checklist with the RECORD extension

**Manuscript:** Routine health information system data quality in a hill and mountain district of Nepal: a cross-sectional desk review of four years of DHIS2 data from Sankhuwasabha, 2021-2025

The RECORD statement (REporting of studies Conducted using Observational Routinely-collected health Data) extends the STROBE checklist for studies using routinely collected data (Benchimol EI, et al. PLOS Medicine 2015;12(10):e1001885). Item descriptions below are condensed; the full wording is at www.record-statement.org.

Both reporting statements are cited in the manuscript itself (Methods, "Analysis": STROBE reference 15, von Elm E, et al. Lancet 2007;370(9596):1453-7; RECORD reference 16), and that paragraph points to this completed checklist as S1 Checklist.

#### STROBE items with RECORD extensions

| **Item** | **Checklist item (condensed)** | **Location in manuscript** |
|---|---|---|
| **1. Title and abstract** | | |
| 1(a) | Indicate the study design in the title or abstract | Title ("cross-sectional desk review"); Abstract, Methods ("desk review") |
| 1(b) | Informative and balanced summary in the abstract | Abstract (structured) |
| RECORD 1.1 | Type of data used specified in title or abstract; name the databases | Title and Abstract ("DHIS2 data"; "aggregated DHIS2 data") |
| RECORD 1.2 | Geographic region and timeframe in title or abstract | Title ("Sankhuwasabha", "Nepal", "four years", "2021-2025"); Abstract ("fiscal year 2024/25 against three preceding years") |
| RECORD 1.3 | If databases were linked, state this in title or abstract | Not applicable (no record linkage; single-platform aggregated data) |
| **2. Background/rationale** | Scientific background and rationale | Introduction, paragraphs 1-4 (geographic gap; validation-rule gap) |
| **3. Objectives** | State specific objectives | Introduction, final paragraph (two stated aims) |
| **4. Study design** | Present key elements of study | Methods, "Study design and setting" |

| | | |
|---|---|---|
| | design early | |
| **5. Setting** | Setting, locations, relevant dates (periods of data collection) | Methods, "Study design and setting" (district, 10 palikas; reference year FY 2081/82 = mid-July 2024 to mid-July 2025; comparison years FY 2078/79-2080/81 = 2021/22-2023/24, mid-July 2021 to mid-July 2024; Bikram Sambat calendar explained and Gregorian equivalents given throughout); "Data source and extraction" (extraction dates: reporting rates 24 June 2026, service volumes 1 July 2026) |
| **6. Participants** | Eligibility criteria, sources and selection | Methods, "Study design and setting" (census: all 132 DHIS2-reporting facilities; no sampling) |
| RECORD 6.1 | Methods of study population selection (codes/algorithms) listed in detail | Methods, "Data source and extraction" (7 named datasets, all reporting units); S1 Table (exact element selection) |
| RECORD 6.2 | Validation studies of the codes/algorithms used to select the population | Not applicable (no case-finding algorithm; all reporting units of named datasets included). Element names verified against the district DHIS2 data dictionary (Methods, "Tracer indicators"; S1 Table) |
| RECORD 6.3 | If linkage was used, consider a flow diagram of the linkage process | Not applicable (no linkage) |
| **7. Variables** | Define all outcomes, exposures, predictors | Methods, "Tracer indicators" (including a worked account of how derived indicators are summed from their component DHIS2 elements, using institutional delivery as the example) and "Data-quality metrics and benchmarks" |
| RECORD 7.1 | Complete list of codes and | S1 Table (exact DHIS2 element names |

| | | |
|---|---|---|
| | algorithms used to classify exposures/outcomes | and derivation of every summed indicator); S2 Table (pair definitions and rule classification) |
| **8. Data sources/measurement** | Sources of data and methods of assessment for each variable | Methods, "Data source and extraction" (reporting-rate and service-volume exports; custodian extraction) |
| **9. Bias** | Efforts to address potential sources of bias | Methods, "Analysis" (pre-specified benchmarks and pair classification; advance flagging of small-count indicators); Methods, "Classification of related-indicator pairs" (pairs classified as rule-governed or free before any metric was computed, and rule-governed pairs set aside before consistency conclusions are drawn) |
| **10. Study size** | Explain how the study size was arrived at | Methods, "Study design and setting" (census of all 132 facilities; no sampling) |
| **11. Quantitative variables** | How quantitative variables were handled; groupings and why | Methods, "Data-quality metrics and benchmarks" (metrics, thresholds, benchmarks and their sources) |
| **12. Statistical methods** | Describe all statistical methods, incl. subgroups and missing data | Methods, "Data-quality metrics and benchmarks" and "Analysis" (all metrics descriptive; district and palika subgroup resolution; blanks treated as zero; no imputation; no gaps found in reference-year series; software named with versions (Python 3.12.0, xlrd 2.0.2, openpyxl 3.1.5); pipeline stated to be deterministic with no stochastic step) |
| RECORD 12.1 | Extent of investigators' access to the database population | Methods, "Data source and extraction" (Health Office is the district data custodian with routine DHIS2 access); "Competing interests" |

| | | |
|---|---|---|
| RECORD 12.2 | Information on data cleaning methods | Methods, "Analysis" (exports analysed as extracted; no imputation; blank-cell rule; completeness of monthly series checked) |
| RECORD 12.3 | State whether the study included data linkage; linkage methods and quality | Not applicable (no linkage; single aggregated platform) |
| **13. Participants (results)** | Numbers at each stage; reasons for non-participation | Results, "Completeness and timeliness" (7 datasets x 11 reporting units; 70 palika-dataset combinations; no unit excluded) |
| RECORD 13.1 | Detail selection of included persons/units, incl. filtering on data quality or availability | Results, "Completeness and timeliness" (no filtering applied; all reporting units retained); Methods, "Tracer indicators" (pre-specified OPD asymmetry and its reason) |
| **14. Descriptive data** | Characteristics of study participants/units | Methods, "Study design and setting" (district population, terrain, facility count); Results, Table 1 and service-volume paragraph |
| **15. Outcome data** | Report numbers of outcome events or summary measures | Results, Tables 1-3, Fig 1 and text (all metric values); S3 Table (numerators and denominators behind every palika ratio, and the district aggregation check) |
| **16. Main results** | Unadjusted estimates and their precision, as applicable | Results (descriptive metrics against pre-specified benchmarks; survey denominators given for NDHS estimates in Table 3 note) |
| **17. Other analyses** | Report other analyses done | Results, "Consistency over time, outliers, and the composite report card"; four-year trend analysis (Table 2 and Fig 1); "External plausibility check" (Table 3) |
| **18. Key results** | Summarise key results with reference to objectives | Discussion, "Principal findings" |

| | | |
|---|---|---|
| **19. Limitations** | Discuss limitations, incl. sources of potential bias | Discussion, "Strengths and limitations" |
| RECORD 19.1 | Discuss implications of using data not created to answer the research question | Discussion, "Strengths and limitations" (palika-level resolution of a facility-based source; no source-document verification; the rule document records which relationships carry a rule but not the enforcement mode or enablement date; proposed mechanisms for the immunization inversion stated as candidates, not findings) and "Validation rules and the consistency toolkit" (platform configuration shaping the data) |
| **20. Interpretation** | Cautious overall interpretation | Discussion, "Principal findings"; Conclusions |
| **21. Generalisability** | Discuss generalisability | Discussion, "Strengths and limitations" (palika findings district-specific; methodological point general to rule-enabled DHIS2) |
| **22. Funding** | Source of funding and role of funders | "Funding": no specific funding |
| RECORD 22.1 | How to access supplemental information: protocol, raw data, programming code | "Data availability"; S1 Text (analysis code openly available, https://doi.org/10.5281/zenodo.21211933; aggregated exports and analysis workbook deposited on Zenodo, https://doi.org/10.5281/zenodo.23149871) |
